# Beyond Words: Natural Language Sampling of Lexical and Non-Word Vocalizations in Preschool Children with Autism, Down Syndrome, and Fragile X Syndrome

**DOI:** 10.64898/2026.09.21.26363560

**Authors:** Anna Adekogbe, Antuan Tran, Gabriela Miller, Margaret Norberg, Alexis Monk, Tanisha Chanda, Xinran Hou, Katherine Palowski, Nicole Baumer, Kristina T. Johnson, Carol Wilkinson

## Abstract

**Purpose:** This study examined whether word-based and non-word natural language sampling (NLS) measures complement standardized language scores in characterizing expressive communication among preschool children with autism with language impairment (AUT-LI), Down syndrome (DS), and fragile X syndrome (FXS).

**Method:** Participants included 82 preschool children with AUT-LI (*n* = 32), DS (*n* = 34), or FXS (*n* = 16). Expressive communication was characterized using the Preschool Language Scales and 10-minute natural language samples collected during a standardized play-based protocol. NLS measures included traditional word-based metrics, such as lexical diversity, intelligibility, and word approximations, as well as tiered non-word vocalization measures, including non-speech, single-phoneme, and multi-phoneme vocalizations. Analyses examined age associations, diagnostic group differences, between group differences in variability, and exploratory communication profiles derived using principal component analysis and partitioning around medoids clustering.

**Results:** Diagnostic groups did not significantly differ on standardized expressive language scores. Children with FXS showed higher performance than the AUT-LI and DS groups on several word-based NLS measures, whereas non-word vocalization measures primarily revealed differences in within-group variability. Word-based NLS measures were consistently associated with standardized expressive language skills, while associations for non-word vocalizations varied by vocalizations type. Cluster-validity indices factored a two-cluster solution. A hypothesis-informed three cluster solution that included non-word vocalizations suggested Higher Language, Lower Output, and High Output-Low Structure profiles.

**Conclusions:** Standardized and NLS measures provide complementary descriptions of expressive communication in children with neurodevelopmental disorders. Non-word vocalization coding may be especially useful for describing clinical meaningful variation among children with limited expressive language.

## Introduction

Children with neurodevelopmental disorders (NDDs) exhibit highly diverse developmental trajectories, particularly in the domain of language (Lord et al., 2020; Marschik et al., 2022; Tager-Flusberg & Kasari, 2013). While some individuals experience only mild challenges, a significant subgroup of individuals remain minimally verbal or fail to acquire functional multiword speech despite receiving years of intervention (Kasari et al., 2013; Tager-Flusberg & Kasari, 2013; Thurm et al., 2015). Characterizing these language profiles becomes increasingly difficult with co-occurring intellectual disability (ID), as clinical nuances are frequently lost when assessment tools created and normed for more verbal populations are applied to individuals with significant impairments (Abbeduto et al., 2014; Hessl et al., 2009; Kasari et al., 2013). This often results in a loss of symptom detail critical for phenotype differentiation (Abbeduto et al., 2014; Hamrick et al., 2025). To address these disparities, consistent and nuanced approaches for language assessment are required both within and across groups for meaningful comparison (Abbeduto et al., 2014; Brady et al., 2016; Kasari et al., 2013). Accurate characterization of language abilities is therefore critical for monitoring progress, guiding intervention, and supporting individualized educational and life-skill planning (Brady et al., 2021; Hessl et al., 2009; Kasari et al., 2013). Without sensitive and developmentally appropriate tools, children risk delayed services or placement in suboptimal treatment pathways (Belardi et al., 2017; Brady et al., 2021).

### Limitations of standardized assessments

Standardized psychometric tests are the most widely used tools for evaluating expressive language, but they often fail to provide a complete picture of communication for those with significant impairments (Condouris et al., 2003; Kasari et al., 2013; Spaulding et al., 2006). These assessments are also vulnerable to floor effects, in which many children receive the lowest possible score or cluster at the bottom of the scoring range, limiting the measure’s ability to distinguish meaningful differences among children with limited language ability (Barokova et al., 2021; Finestack & Abbeduto, 2010; Hessl et al., 2009). In addition, children with lower language levels are frequently excluded from studies, leading to a lack of empirical knowledge regarding the “neglected end” of the spectrum (Kasari et al., 2013; Tager-Flusberg & Kasari, 2013). Standard assessments may also lack sensitivity to detect small but meaningful developmental changes or the influence of pragmatic factors on a child’s real-time performance (Barokova & Tager-Flusberg, 2018; Condouris et al., 2003). Finally, standardized testing environments may underestimate a child’s language and communication ability, as the rigid requirements for rapport and compliance can interfere with the performance of children with NDDs (Barokova et al., 2020; Tager-Flusberg & Kasari, 2013).

### A Multi-method framework for language assessment

Because the level of language impairment varies greatly among NDDs, previous literature advocates for a multi-method approach to characterizing language in children with limited expressive language (Barokova & Tager-Flusberg, 2018; Brady et al., 2016; Kasari et al., 2013; Tager-Flusberg & Kasari, 2013). There is evidence that a framework combining standardized metrics and systematic natural language sampling (NLS) may better capture the communicative repertoire of children with NDDs (Gary & Wallace, 2020; Pavelko et al., 2016; Peña et al., 2014; Tager-Flusberg et al., 2009). Importantly, NLS captures real-time functional communication in naturalistic settings, and rate-based NLS measures provide detailed insight into communication differences across NDD groups (Plate, 2025; La Valle et al., 2020; Woynaroski et al., 2016).

At the lexical level, measures derived from NLS sessions, such as Mean Length of Utterance (MLU) and Number of Different Words (NDW), provide sensitive indices of grammatical complexity and lexical diversity that are often more sensitive to language differences than standardized scores alone (Condouris et al., 2003; Heilmann & Miller, 2023). Second, NLS captures critical data on speech intelligibility, word approximations, and gesture use to supplement speech (La Valle et al., 2024). Despite its utility, NLS remains underutilized in part due to a lack of standardized, feasible coding schemes for early vocalizations in minimally speaking populations, particularly in clinical research involving children with severe expressive language impairment (Barokova et al., 2020; Gary & Wallace, 2020; Costanza-Smith, 2010; Kasari et al., 2013).

Importantly, NLS metrics can also be used to capture the infrastructural building blocks of speech, known as protophones, which serve as the essential precursors to formal language development (Johnson et al., 2023; Narain et al., 2020a). Protophones are volitional, speech-like precursors that exhibit functional flexibility across emotional states and social contexts (Jhang & Oller, 2017; Marschik et al., 2022; Johnson et al., 2023; Narain et al., 2020b). To map a child’s progression toward spoken language, vocalizations can be grouped into three structural classes based on phonetic complexity. Vocalizations at any level may be socially directed, exploratory, or stereotyped depending on the context (Oller et al., 2013):

1. Nonspeech-like vocalizations: Cries and other non-segmental sounds such as grunts, squeals, growls, and idiosyncratic vocalizations (Marschik et al., 2022).
2. Pre-canonical (single-phoneme) vocalizations: isolated consonant-like or vowel-like sounds (Lynch et al., 1995; Marschik et al., 2022).
3. Canonical babbling (multi-phoneme) vocalizations: well-formed syllables with rapid, adult-like timing (Belardi et al., 2017; Marschik et al., 2022; Patten et al., 2014).

Together, these structural levels provide clinically relevant information about early vocal development. Nonspeech-like vocalizations are a common and accessible form of communication for children and individuals with NDDs, particularly for those with limited expressive language (Narain et al., 2020a). Pre-canonical vocalizations may reflect a transition toward canonical babbling and emerging speech-motor control needed for later speech development (Long & Hustad, 2023; Iverson & Wozniak, 2007). Canonical syllables are the constituents of real speech, identifying delays at this level is a predictor of later language impairment across NDD groups (Nathani et al., 2006; Oller & Kent, 2001).

### Disorder-specific communication profiles

Various NDDs exhibit differing vocalization frequency, phonetic complexity, and the achievement of prelinguistic milestones (Belardi et al., 2017; Fidler, 2005; Marschik et al., 2022; Roche et al., 2018). In this study, we assess three NDDs: Autism, Fragile X syndrome (FXS), and Down Syndrome (DS) because of their overlapping yet distinct communication presentation and needs (Abbeduto et al., 2014; Kaufmann et al., 2017).

Autism is characterized by qualitative impairments in social communication and reciprocity (Lord et al., 2020; Tager-Flusberg et al., 2005). While these core difficulties are universal, linguistic profiles across the spectrum are highly diverse; some children with autism develop typical verbal speech, while others present with co-occurring language impairment. It is estimated that 30% of children with autism remain minimally speaking or nonverbal, and do not develop functional spoken language even after receiving years of intensive intervention (Brady et al., 2021; Tager-Flusberg & Kasari, 2013; Thurm et al., 2015; Luyster et al., 2008). Language acquisition is often delayed, with first words appearing at an average of 38 months (Eigsti et al., 2011; Maes et al., 2022; Tager-Flusberg et al., 2005). Early markers of autism include lower rates of volubility (the frequency of vocalization) and lower canonical babbling ratios (Maes et al., 2022; Patten et al., 2014). These deficits may be linked to a diminished social feedback loop between the child and caregiver, which limits the child’s opportunities to hear and produce speech-like sounds (Patten et al., 2014; Warlaumont et al., 2014).

FXS is the leading inherited cause of intellectual disability (CDC, 2025; Finestack et al., 2009; Hessl et al., 2009) and is characterized by significant delays in prelinguistic milestones (Belardi et al., 2017; Finestack et al., 2009). Infants with FXS are significantly less likely to reach the canonical babbling stage by 12 months compared to typically developing peers (Belardi et al., 2017; Roche et al., 2018). Their communication profiles often feature repetitive language or perseveration and significant difficulties in sequential processing and working memory (Abbeduto et al., 2014; Finestack et al., 2009). While the prevalence estimates for autism in FXS vary across studies based on diagnostic tools and assessment methods, the rate of co-occurrence remains consistently high, with literature indicating 46-54% males and 16-20% females meeting criteria (CDC, 2025; Roberts et al., 2020; Kaufmann et al., 2017).

DS is one of the most leading causes of intellectual disability and is associated with substantial language and communication challenges (CDC, 2026). Although both receptive and expressive language impairments are common, expressive language is often more severely affected than receptive language (Chapman, 2006; Fidler, 2005; Finestack & Abbeduto, 2010). Expressive challenges in DS include difficulties with speech production, grammar, syntax, and intelligibility, with reduced intelligibility partly reflecting oro-motor and anatomical differences. Children with DS often produce shorter, less complex utterances than language-matched peers, highlighting syntax as a persistent area of difficulty (Chapman, 2006; Finestack & Abbeduto, 2010; Hamrick et al., 2025). However, studies have shown that communicative attempts and volubility rates (vocalizations per minute) may be comparable to typically developing peers (Belardi et al., 2017). Substantial variability exists within DS communication profiles (Thurman et al., 2022), underscoring the need for measures that capture both language structure and vocal output.

### Current study

This study examines how standardized language assessments and NLS measures at both lexical and vocalization levels capture expressive communication heterogeneity across three overlapping yet distinct neurodevelopmental groups: autism with language impairment (AUT-LI), DS, and FXS. The present study leverages a cross-sectional cohort from the Brain Indicators of Developmental Growth (BRIDGE) study to determine if integrating traditional NLS metrics with non-word measures enhances the differentiation of language phenotypes. We sought to answer the following research questions:

1. To what extent do standardized language assessments and word-based NLS metrics capture variability in communication profiles across AUT-LI, FXS, and DS? We hypothesize that while standardized scores will show comparable levels of impairment across groups, both rate-based NLS measures (e.g., intelligible words per minute) and lexical diversity measures (NDW; number of different words) will reveal significant group differences and capture variability that traditional norm-referenced metrics do not capture.
2. Do novel non-word NLS measures, including both non-speech-like vocalizations and speech-like vocalizations, provide additional information about heterogeneity in expressive communication? We predict that the rate of novel non-word NLS measures (non-speech like vocalizations per minute, single-phoneme vocalizations per minute, and multi-phoneme vocalization per minute) will provide unique information about expressive communication not captured by standardized language measures or traditional word-based NLS metrics.

## Method

Institutional Review Board approval for the BRIDGE study was obtained from Boston Children’s Hospital (IRB-P00034676). Legal guardians of all participants provided written informed consent prior to participation.

### Participants and recruitment

Participants (n = 82) were drawn from a longitudinal investigation studying neurodevelopmental change in language across multiple disorders: AUT-LI (n = 32), FXS (n = 16), and DS (n = 34). Study participants, aged 24 to 68 months, were seen two times roughly 1 year apart (see Table 1 for full sample demographics). For this analysis, we utilized only Time Point 1 data for all metrics, with two exceptions where data from both timepoints were used: (1) when assessing the relationship between age and standardized scores, incorporating Time Point 2 data where available, and (2) when assessing change in PLS expressive communication GSV between Time Point 1 and Time Point 2 among participants with available longitudinal data (n = 51; AUT-LI: n = 18; DS: n = 26; FXS: n = 7).

**Table 1.** Sample Characteristics.

| **Table 1. Sample Characteristics** |  |  |  |  |
| --- | --- | --- | --- | --- |
|  | **Full Sample**<br>**N = 82** | **ASD-LI**<br>**N = 32** | **DS**<br>**N = 34** | **FXS**<br>**N = 16** |
| **Chronological age (months)** | 45.9 (13.4) | 50.4 (10.9) | 42.1 (14.5) | 44.6 (13.5) |
| **Child Sex, *n* (%)** |  |  |  |  |
| Male | 67 (81.5) | 31 (96.9) | 20 (58.8) | 16 (100.0) |
| Female | 15 (18.5) | 1 (3.1) | 14 (41.2) | 0 (0.0) |
| **Child Race, *n* (%)** |  |  |  |  |
| White | 58 (70.7) | 18 (56.2) | 28 (82.4) | 12 (75.0) |
| Black or African American | 10 (12.2) | 8 (25.0) | 2 (5.9) | 0 (0.0) |
| Mixed | 6 (7.3) | 2 (6.2) | 1 (2.9) | 3 (18.8) |
| Asian | 5 (6.1) | 4 (12.5) | 1 (2.9) | 0 (0.0) |
| Other | 2 (2.4) | 0 (0.0) | 1 (2.9) | 1 (6.2) |
| Not Reported | 1 (1.2) | 0 (0.0) | 1 (2.9) | 0 (0.0) |
| **Ethnicity, *n* (%)** |  |  |  |  |
| Not Hispanic | 70 (85.4) | 27 (84.4) | 30 (88.2) | 13 (81.2) |
| Hispanic | 11 (13.4) | 5 (15.6) | 3 (8.8) | 3 (18.8) |
| Not Reported | 1 (1.2) | 0 (0.0) | 1 (2.9) | 0 (0.0) |
| **Parental Education, *n* (%)** |  |  |  |  |
| < 4-year college degree | 12 (14.7) | 6 (18.7) | 5 (14.7) | 1 (6.2) |
| 4-year college degree | 28 (34.1) | 11 (34.4) | 9 (26.5) | 8 (50.0) |
| > 4-year college degree | 42 (51.8) | 15 (46.8) | 20 (58.8) | 7 (43.7) |
| **Household Income, *n* (%)** |  |  |  |  |
| < \$40,000 | 2 (2.4) | 1 (3.1) | 1 (2.9) | 0 (0.0) |
| \$40,000 - \$70,000 | 14 (17.1) | 6 (18.8) | 7 (20.6) | 1 (6.2) |
| \$70,000 - \$100,000 | 14 (17.1) | 5 (15.6) | 6 (17.6) | 3 (20.0) |
| \$100,000 - \$140,000 | 17 (20.7) | 6 (18.8) | 6 (17.6) | 5 (31.2) |
| > \$140,000 | 34 (42.0) | 14 (43.8) | 13 (38.2) | 7 (43.8) |
| Not Reported | 1 (1.2) | 0 (0.0) | 1 (2.9) | 0 (0.0) |

Note. Means and standard deviations are reported for chronological age.

Participants in the syndrome groups required a confirmed genetic diagnosis of trisomy 21 or FXS with full mutation. We acknowledge that a subset of participants in the DS and FXS groups also met clinical criteria for an autism diagnosis (DS: n = 1; FXS: n = 7). Due to sample size considerations, participants in these groups were classified according to their primary genetic diagnosis. Children in the AUT-LI group required a clinical diagnosis of autism spectrum disorder from a licensed professional. To ensure phenotypic comparability with the naturally occurring communication profiles of children with DS and FXS, inclusion criteria for the AUT-LI group required co-occurring language impairment, defined as a receptive and/or expressive standard score of 85 or below on the Preschool Language Scales–Fifth Edition (PLS; Zimmerman et al., 2011). This entry criterion allowed for a cohort in which traditional assessments are most prone to floor effects and likely to mask meaningful communicative variability.

All participants were required to have at least 75% daily exposure to English. Exclusion criteria included a history of significant prematurity (< 34 weeks), known birth trauma, unstable seizure disorders, or uncorrected hearing/vision impairments. To prevent interference with transcription, technical exclusions were made for participants who completed the NLS protocol while wearing face masks (n=7) during COVID health and safety procedures. No minimum utterance threshold was applied; participants were included regardless of vocalization rate to be inclusive of communication profiles and to avoid biasing toward more verbal children.

## Measures

### Preschool Language Scales - 5^th^ Edition (PLS)

The PLS was administered to evaluate expressive communication and auditory comprehension across vocabulary, grammar, and prelinguistic domains (Zimmerman et al., 2011). Analysis focused on scores from the expressive communication subdomain, including the expressive communication standard scores (PLS SS), growth scale values (PLS GSV), age equivalencies (PLS AE). PLS developmental quotient scores (PLS DQ), calculated by averaging age-equivalents across subdomains, dividing by actual age in months and multiplying by 100, were used for characterization and longitudinal growth analyses.

### Mullen Scales of Early Learning (MSEL)

The MSEL is a standardized developmental assessment designed to evaluate cognitive and motor functioning in young children (Mullen, 1995). In the current study, analyses focused on the nonverbal developmental quotient (NVDQ) derived from the visual reception and fine motor subscales of the MSEL. This measure was used to characterize participants’ nonverbal developmental abilities and to contextualize expressive language performance relative to nonverbal cognitive skills across diagnostic groups.

### Eliciting Language Samples for Analysis Toddler Edition (ELSA-T)

Vocalizations were elicited using the (ELSA-T), a play-based protocol designed to promote spontaneous expressive communication (Barokova et al., 2021). ELSA-T is structured to accommodate children with diverse language levels and behavioral challenges, ensuring high-quality capture of language behaviors in a standardized setting. It uses conversation, play, and narrative elicitation contexts to create multiple diverse opportunities for a child to produce spontaneous language. Some of these activities include hide and seek and looking at books together. Administration time typically lasts 15-25 minutes, depending on developmental ability and engagement. For analytic consistency, the first 10 minutes of ELSA were transcribed. Periods of caregiver interruption or off-task exchanges were excluded. The resulting average transcript length was 9.9 minutes (SD = 0.24; range: 8.25–10.45), providing participants with equivalent opportunities for language production within a standardized context.

### Transcription scheme and reliability

Language samples were analyzed using a novel tiered transcription protocol that uses the Systematic Analysis of Language Transcripts (SALT) format (Miller & Iglesias, 2012), while incorporating additional phonetic features designed to characterize communication in children with limited expressive language. A novel contribution of this study is the use of a tiered coding framework that extends beyond transcribable words, to capture non-word vocalizations. Specifically, the framework includes customized categorization of speech productions like non-speech, single-phoneme and multi-phoneme vocalizations (Figure 1). Because standard SALT software is not designed to collect these prelinguistic features and defaults to including only complete and intelligible utterances in analysis, a custom Python script was developed to analyze and derive NLS measures. Both the analysis script and detailed description of the protocol are available on GitHub (see Data Availability Statement).

**Figure 1.**
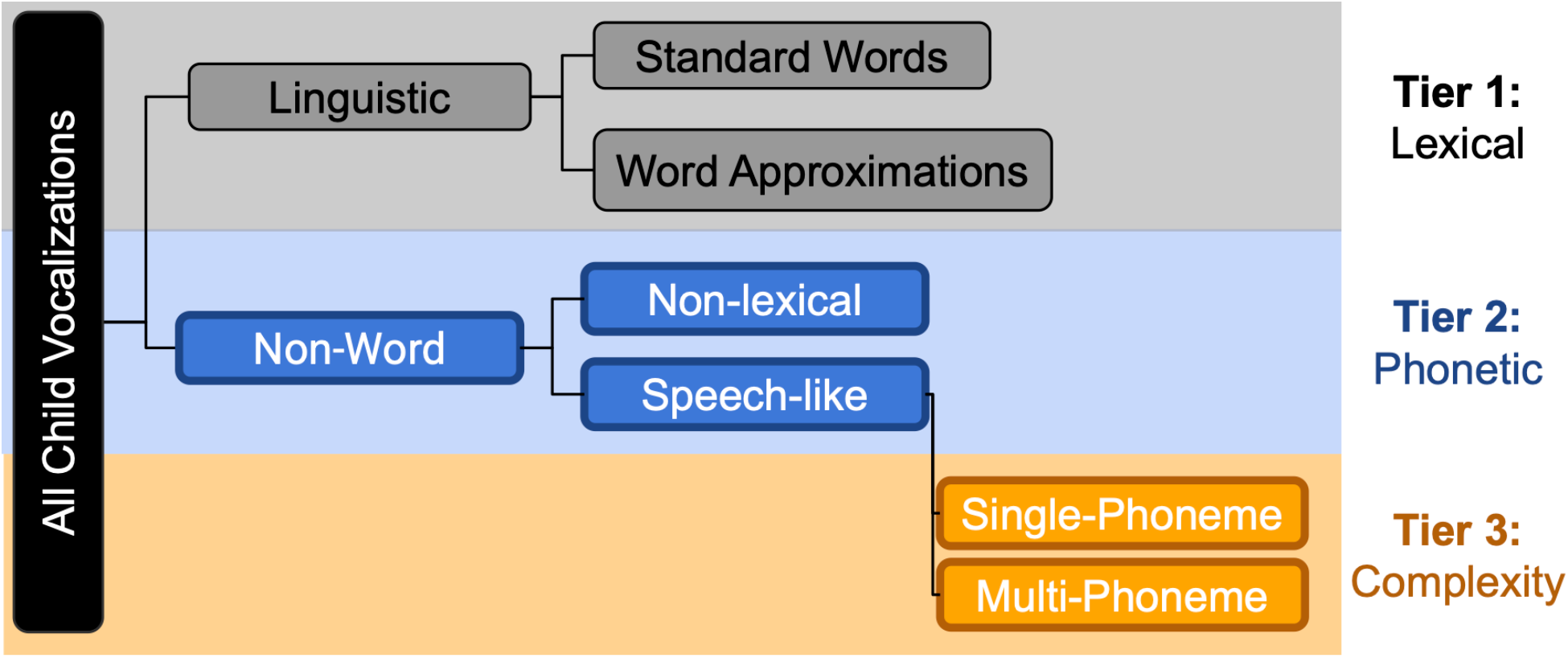
Tiered Transcription Coding Schematic.

Transcribers were trained using SALT modules and supervised practice samples until they achieved a minimum of 80% agreement with master training transcripts. A sequential review process was used to support transcription quality. Research assistants first completed standard SALT transcription, including transcription of standard words and phrases and identification of unintelligible productions, and marked productions they identified as potential word approximations. Each first-pass transcript was subsequently reviewed against the corresponding NLS video by the lead graduate-level transcriber, who verified the initial transcription and reviewed and confirmed word-approximations classifications. Disagreements were resolved through group adjudication and consensus. Non-word vocalization categories were coded during higher-tier review by the lead graduate-student transcriber according to the study coding protocol.

As a transcript quality-control check, a random subset of 16 transcripts (approximately 20% of the sample) was selected to evaluate agreement between measures derived from the original first-pass and finalized transcripts. Two-way, absolute-agreement, single-measure intraclass correlation coefficients indicated strong correspondence for percent unintelligible utterances (ICC= .911, 95% CI [.735, .969]), percent response to questions (ICC = .910, 95% CI [.520, .975]), and total utterances (ICC = .995, 95% CI [.985, .998]). As a practical assessment of first-pass transcription quality and the impact of sequential review, median absolute differences between first-pass and finalized transcripts were also calculated. Median differences were 0.91 percentage points for unintelligible utterances, 2.32 percentage points for response to questions, and 1.5 utterances for total utterances. These comparisons reflect changes following sequential review rather than independent interrater reliability for the non-word coding categories.

#### Language Measure Definitions

For intelligible speech, utterance segmentation followed standard SALT conventions using communication units (C-units), defined as an independent clause with its modifiers. Each C-unit included one main clause and any attached subordinate clauses and could not be further divided without changing its essential meaning. For non-word and nonspeech-like vocalizations, boundaries were primarily determined using a breath-group criterion, with a new vocalization event beginning after an audible breath or a pause of 1 second or more. When distinct vocalization types occurred within the same breath group, such as a speech-like vocalization followed by a squeal, these were coded as separate vocalization events to preserve distinctions among vocalization categories.

The following metrics were extracted from transcripts using a custom Python script:

### Word-based Language Measures

#### Mean Length of Utterance in Words (MLUw)

The average number of intelligible words per child utterance. MLUw was calculated by summing the number of intelligible words across child utterances and dividing by the total number of utterances. Unintelligible utterances were not counted as words, but utterances with no intelligible words remained in the denominator and contributed a value of zero. For example, a child with 10 single-word utterances and 10 utterances with no intelligible words would have an MLUw of 0.50, whereas a child with 20 intelligible single-word utterances would receive an MLUw of 1.00.

#### Number of Different Words (NDW)

A measure of lexical diversity, summarizing the rate of unique intelligible words produced. Both word approximations and imitative productions were included in the NDW calculation.

#### Intelligibility

The proportion (%) of total child utterances that were clearly understandable and transcribable word forms.

#### Word Approximations (WA)

The rate per minute of phonologically related, non-standard word forms consistently used to represent a target word. WAs were treated as intelligible speech and were included in all lexical and grammatical calculations (e.g., MLUw, NDW, and intelligibility).

### Non-Word Vocalization Categories

Following the tiered protocol (Figure 1), vocalizations that did not constitute identifiable words were stratified by complexity:

#### Non-Lexical Vocalizations

Vocalizations lacking a transcribable phonetic structure, including grunts, cries, raspberries, and idiosyncratic sounds.

#### Single-Phoneme Vocalizations

Speech-like productions consisting of isolated vowels or consonants.

#### Multi-phoneme Vocalizations

Vocalizations containing consonant-vowel (CV) combinations, including single CV syllables and repeated or varied CV sequences. Productions were coded as multi-phoneme vocalizations regardless of whether they showed rapid, adult-like timing.

All NLS measures (except for MLUw and Intelligibility) were converted into rates by taking each variable and dividing it by duration of the transcript in minutes to account for any difference in total duration.

### Statistical Analyses

#### Aim 1: Age Associations, Phenotypic Characterization, and Variability

To examine the relationship between chronological age and expressive language outcomes, we used linear mixed-effects models for expressive communication standard scores (PLS SS) and growth scale values (PLS GSV). Models included centered age, diagnostic group, and the age × group interaction as fixed effects, with a random intercept for participants to account for repeated observations from children who contributed data at both visits. Available Time point 1 and Time Point 2 observations roughly one -year apart, were included. For NLS metrics, distributions were frequently lower-bounded at zero with substantial floor effects. Associations between chronological age and each NLS outcome were initially examined using Spearman rank correlations because the measures were non-normally distributed. However, the high frequency of zero values resulted in extensive tied ranks and estimated p-values, limiting the interpretability of the rank-based analyses. Therefore, left-censored Tobit regression models were used as the primary analytic approach. Models treated zero as the lower bound of the observed outcome distribution and examined associations between age and each NLS measure, with the diagnostic group included as a covariate.

To characterize the clinical phenotypes, group differences in central tendency for standardized and NLS measures were evaluated using Kruskal–Wallis tests. For measures with significant omnibus Kruskal-Wallis test, post-hoc Dunn tests were used to identify pairwise group differences, with false discovery rate correction applied across pairwise comparison within each measure. To evaluate whether variability differed across diagnostic groups, Brown-Forsythe tests were used to compare group variances for each standardized language and NLS measure. For measures with significant Brown-Forsythe tests, we examined group-specific dispersion using descriptive stats. Finally, the relationship between standardized expressive scores and real-time vocalization rates was assessed using Spearman rank-order correlations.

#### Aim 2: Exploratory Characterization of Communication Profiles

To explore whether inclusion of non-word vocalization measures revealed additional structure in expressive communication profiles, we conducted an exploratory two-stage clustering analysis using principal component analysis (PCA) followed by partitioning around medoids (PAM) clustering. Before conducting PCA, we examined Spearman correlations among candidate NLS variables to identify highly redundant measures. Variables with very high correlations (ρ ≥ .90) were not entered together into the PCA. Among the highly correlated word-based measures, percent intelligible speech was retained because of its clinical interpretability as an index of functional speech clarity, while MLUw, NDW, and intelligible words were excluded from the clustering models. Prior to PCA, all retained clustering variables were z-scored by subtracting the sample mean and dividing by the sample standard deviation, placing all measures on a common scale. All participants included in the clustering analysis had complete data for the variables used in the PCA/PAM models (n = 82).

PCA was then used as a preprocessing step to reduce dimensionality and summarize shared variance across the retained standardized language and NLS measures before clustering (Zheng et al., 2020). This approach was selected to improve stability of the clustering solution given the modest sample size. Because several NLS variables were floor-heavy and non-normally distributed, PCA was used descriptively as a dimension-reduction tool rather than as a confirmatory latent-variable model. Components were retained until they explained at least 75% of the total variance.

Two clustering models were compared descriptively. Model A included standardized expressive language scores and core word-based NLS measures and retained two principal components. Model B added tiered non-word vocalization rates and retained three principal components. PAM clustering was selected because it is less sensitive to extreme values than centroid-based approaches and was therefore appropriate for exploratory clustering of floor-heavy NLS data. For each model, cluster solutions were evaluated using silhouette width and gap statistics across candidate values of k. Although these indices initially supported a two-cluster solution, a three-cluster solution for Model B was also examined as an exploratory sensitivity analysis based on the a priori hypothesis that children with similar expressive language levels may differ in vocalization complexity and speech output efficiency.

After clusters were identified, clusters were exploratorily compared on external clinical variables to aid interpretation of the cluster structure. For each variable, we tested assumptions of normality. ANOVA with Tukey post-hoc tests was used for variables that were approximately normally distributed, including chronological age and MSEL nonverbal developmental quotient. Kruskal-Wallis tests with pairwise Wilcoxon comparisons were used for variables that were highly non-normal or had small subgroup sizes, including longitudinal growth-rate analyses. Because cluster membership was assigned using Time Point 1 communication features and Time Point 2 data were available for a smaller, diagnostically uneven subset, analyses of monthly GSV change were considered exploratory and interpreted descriptively rather than as adjusted tests of cluster-related growth differences. Fisher’s exact tests were used to examine whether clinical syndrome was associated with cluster membership. When significant associations were observed, standardized residuals were examined to identify which diagnosis-cluster combinations contributed most strongly to the association. All statistical analyses were conducted in Python 3.11.5 {NumPy and pandas}, and in R 4.5.0 {VGAM, lme4, lmerTest, cluster, factoextra, irr, car, FSA, tidyverse, patchwork, scales, corrplot, and ggcorrplot}.

## Results

### Age Associations with Norm-Referenced Scores and Raw Expressive Skill

In typical development, norm-referenced standard scores are expected to remain relatively stable across age. However, using available Time Point 1 and Time Point 2 observations, we observed significant declines in PLS expressive communication SS with increasing age (Figure 2). Linear-mixed effects models indicated a significant negative association with age (β = −.22, *p* = .003; AUT-LI reference), indicating that these cohorts fell behind same-age expectations. While no significant differences in standard scores were observed between diagnostic groups at the mean age (all *p-values* >.39), significant Age × Group interactions demonstrated that these declines relative to age-norms were significantly steeper for both the DS (β = −.38, p < .001) and FXS (β =−0.42, *p* < .001) groups compared to AUT-LI. In contrast, analyses of PLS expressive communication GSV, which reflect skill acquisition independent of age-norms, indicated a significant positive association with age (β = 2.37, *p* < .001; AUT-LI reference). Age × Group interactions indicated that the AUT-LI group showed greater increases in GSV with age compared to DS (β = −1.12, *p* = .005) and FXS (β = −1.41, *p* = .005). These findings indicate that children in all groups acquire expressive skills with age, but at differing rates, resulting in increasingly divergent developmental trajectories.

**Figure 2.**
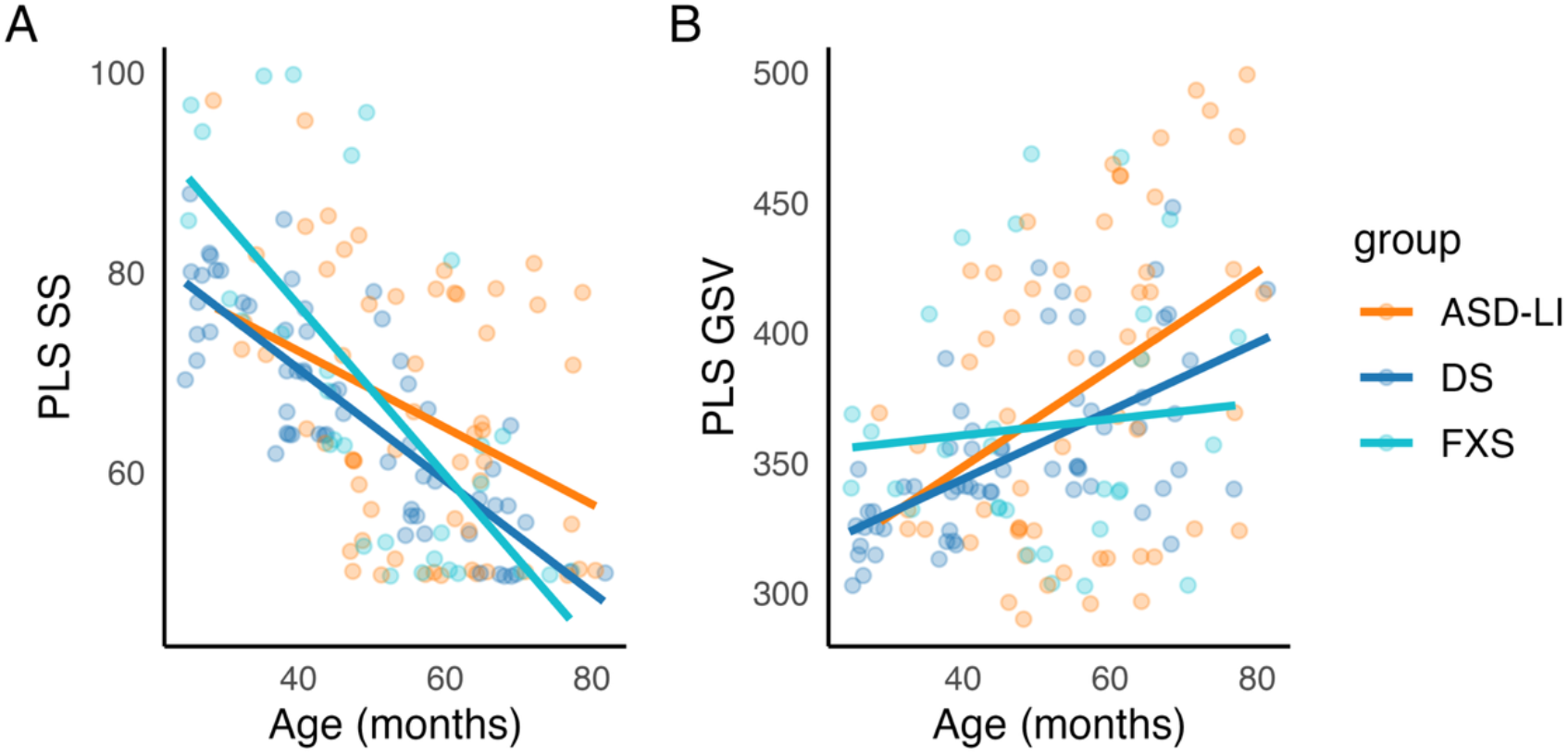
**(A-B).** Standardized expressive language performance trajectories across chronological age by group: Scatterplots with model-estimated linear trajectories for (A) PLS Expressive Communication standard scores (PLS SS) and (B) PLS Expressive Communication Growth Scale Values (PLS GSV).

### Age Associations with NLS Measures

Tobit regression models (Table 2) were used to assess NLS measures association with age across all three clinical groups. Age was significantly and positively associated with MLUw, NDW, rates of intelligible words, and percent intelligible speech. Age was not significantly associated with rates of word approximations, non-lexical vocalizations, or multi-phoneme production. In contrast, age was significantly negatively associated with single-phoneme vocalizations. In contrast to the patterns observed in standardized language performance, Age × Group interactions were not significant for any NLS measure, (*ps* = .064 to .998), suggesting that age-related associations in real-time vocal behaviors did not significantly differ across diagnostic groups.

**Table 2.**
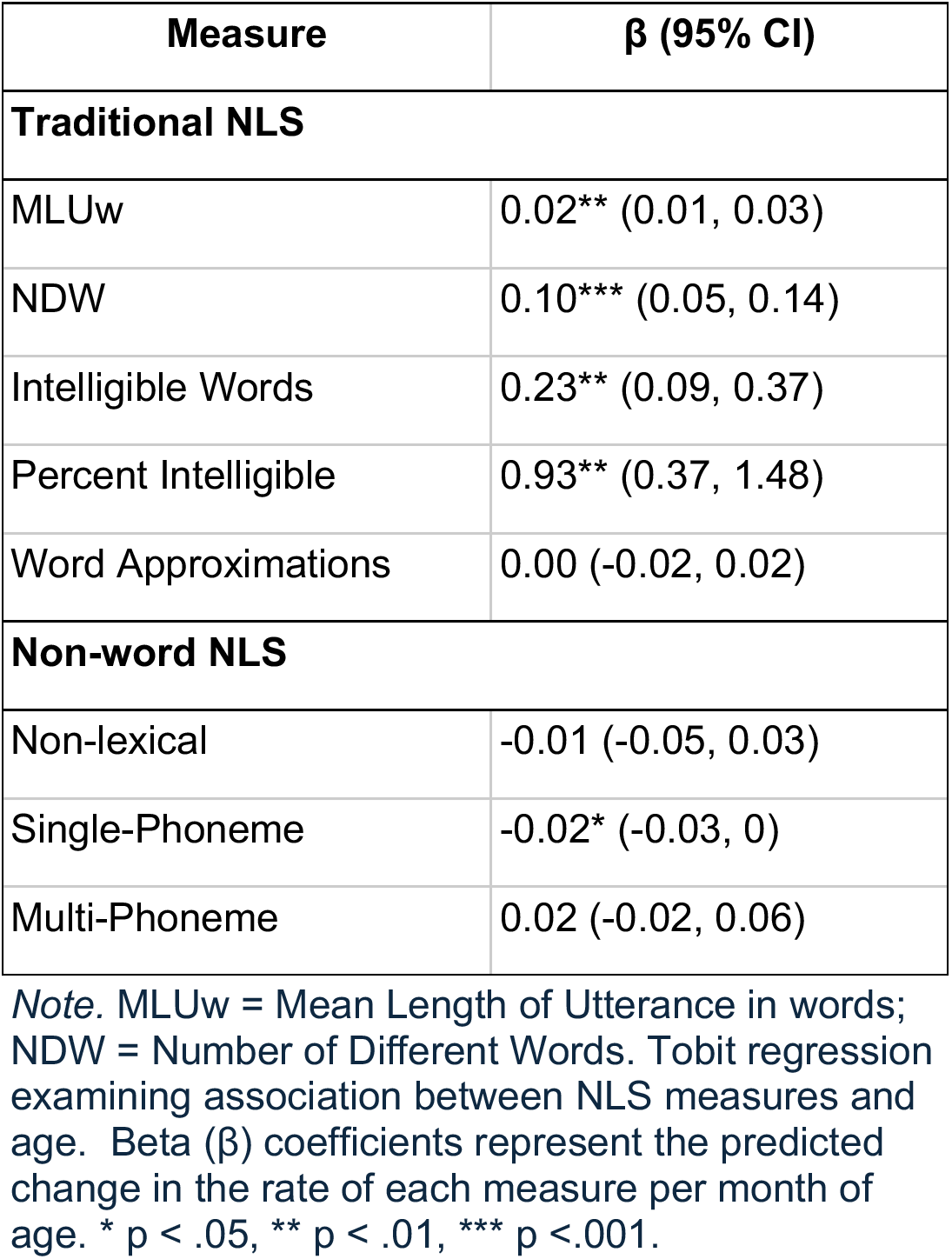
Age Associations with NLS Measures.

### Measure Associations with Standardized Language Skill

To examine the relationship between NLS measures and standardized expressive language scores, Spearman correlations were calculated between NLS metrics and PLS GSV for the full sample and within each diagnostic group. Word-based measures (MLUw, NDW, rate of intelligible words, and percent intelligible) showed a consistent pattern of positive associations across groups (Figure 3). Word approximations showed positive associations with PLS GSV across groups, although the association was weaker in the AUT-LI group.

**Figure 3.**
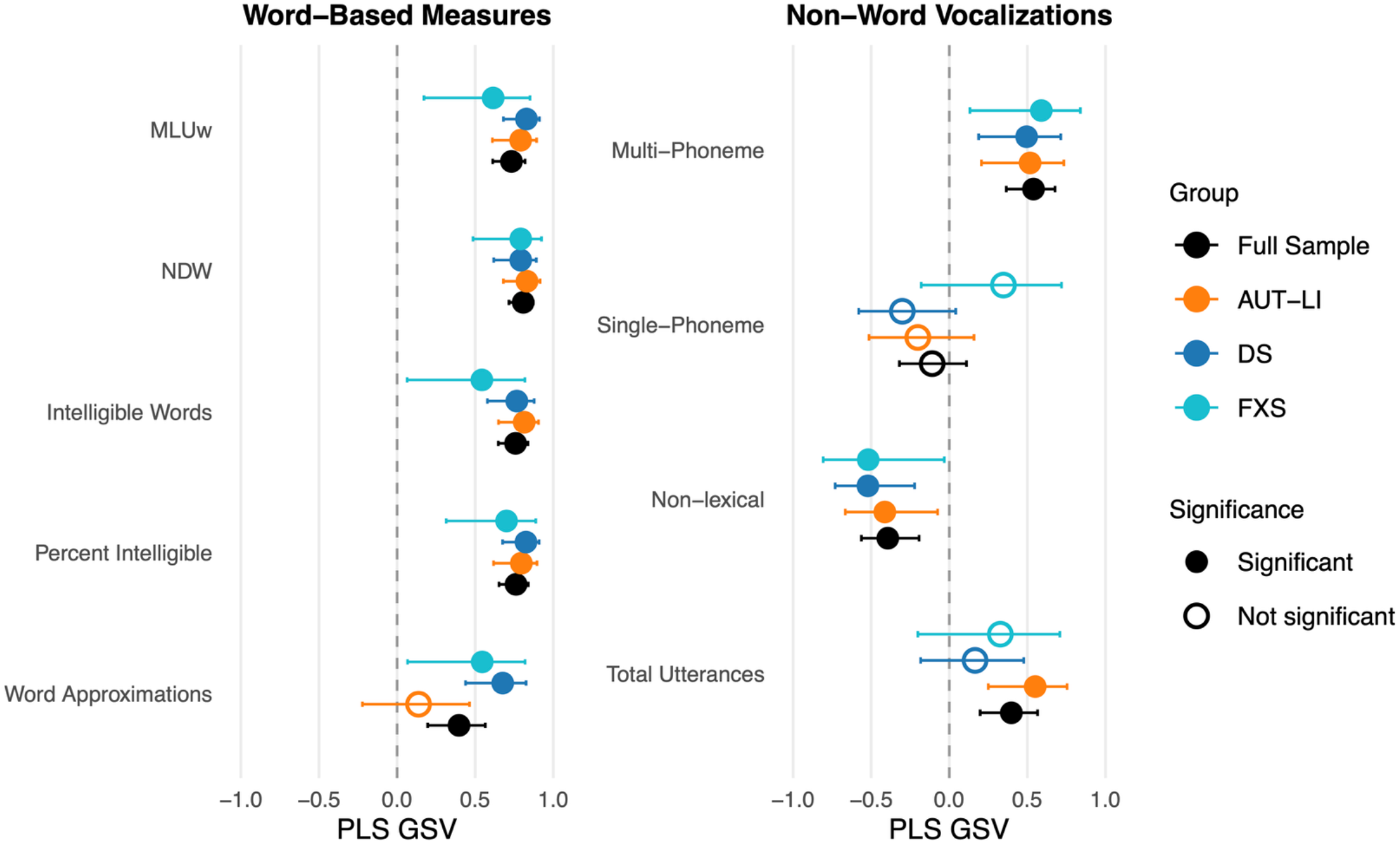
Spearman correlations between PLS Expressive Communication GSV and NLS measures. Error bars represent 95% confidence intervals. Filled points indicate significant correlations (*p* < .05), and open points indicate non-significant correlations.

In contrast, non-word vocalizations showed a more diverse relationship with the PLS GSV. While multi-phoneme vocalizations showed significant positive associations across groups, non-lexical vocalizations (e.g., cries, grunts, squeals) were negatively associated with the PLS GSV across groups. While single-phoneme vocalizations were not statistically significant for any group, the AUT-LI and DS groups showed negative trends, while the FXS group demonstrated a positive trend. Total utterances were positively associated with PLS GSV across groups, with the clearest association observed in the AUT-LI group. Together, these findings suggest that word-based NLS measures showed consistent associations with standardized expressive language scores, while non-word vocalization NLS measures captured more variable patterns of expressive communication across neurodevelopmental conditions.

### Group Comparisons and Communication Heterogeneity

To characterize communication across AUT-LI, FXS, and DS, descriptive statistics for standardized language and cognitive assessments (Table 3) and NLS measures (Table 4) were conducted. Overall, the sample was characterized by limited expressive complexity, marked by short utterances, reduced lexical diversity, and notably low speech intelligibility. Across the clinical cohorts, average intelligibility was very low, ranging from a mean of 15.5% (SD = 20.8) in the DS group to 47.1% (SD= 25.1) in the FXS group. Importantly, while intelligibility was low, the transcription samples were by no means quiet; many children produced frequent vocal output, particularly the FXS group, which averaged 10.4 utterances per minute.

**Table 3.** Descriptives of Age and Standard Language Assessments.

|  | **AUT-LI<sup>1</sup>**<br>n = 32 | **DS<sup>2</sup>**<br>n = 34 | **FXS<sup>3</sup>**<br>n = 16 | **KW**<br>(H) | **BF**<br>(F) |
| --- | --- | --- | --- | --- | --- |
| **Age** (months) | 50.4 (10.9) | 42.1 (14.5) | 44.6 (13.5) | 6.6* (2<1) | 2.4 |
| **PLS** |  |  |  |  |  |
| **DQ** | 47.3 (20.8) | 52.1 (12.7) | 56.5 (24.1) | 2.5 | 5.7** |
| *Expressive* |  |  |  |  |  |
| SS | 64.7 (13.7) | 69.0 (10.8) | 71.6 (17.9) | 3.3 | 2.6 |
| AE | 23.2 (10.5) | 20.9 (4.7) | 23.9 (8.9) | 1.1 | 4.1* |
| GSV | 353.7 (53.4) | 343.6 (25.7) | 358.2 (42.7) | 1.1 | 4.3* |
| *Receptive* |  |  |  |  |  |
| SS | 61.8 (14.4) | 61.9 (9.5) | 64.4 (17.4) | 0.7 | 2.2 |
| AE | 23.3 (12.4) | 20.4 (6.7) | 22.4 (8.2) | 0.6 | 1.9 |
| GSV | 363.5 (57.3) | 354.5 (37.4) | 365.4 (39.0) | 0.6 | 2.8 |
| **MSEL Nonverbal DQ** | 50.9 (18.1) | 50.4 (13.4) | 51.2 (18.1) | 0.04 | 1.9 |

Note. Values are means with standard deviations in parentheses. AE = age equivalents; SS = standard scores; GSV = growth scale values; DQ = developmental quotient. Kruskal-Wallis tests evaluated group differences; significant tests were followed by FDR-corrected Dunn's post-hoc comparisons, with significant pairwise differences shown in parentheses. Pairwise labels are 1 = AUT-LI, 2 = DS, and 3 = FXS. Brown-Forsythe tests evaluated variance differences. \* $p < .05$ , \*\* $p < .01$ , \*\*\* $p < .001$ .

**Table 4.** Descriptives of NLS Measures by Group.

|  | **AUT-LI<sup>1</sup>**<br>n = 32 | **DS<sup>2</sup>**<br>n = 34 | **FXS<sup>3</sup>**<br>n = 16 | **KW**<br>(H) | **Dunn**<br>**Pairs** | **BF**<br>(F) |
| --- | --- | --- | --- | --- | --- | --- |
| **Traditional NLS** |  |  |  |  |  |  |
| MLUw | 0.5 (0.7) | 0.2 (0.3) | 1.0 (0.7) | 14.7*** | 1, 2 < 3 | 4.1* |
| Percent Intelligible | 25.7 (29.8) | 15.3 (20.8) | 47.1 (25.1) | 11.3*** | 1, 2 < 3 | 1.8 |
| Word Approx. | 0.1 (0.2) | 0.1 (0.4) | 0.6 (0.9) | 12.6*** | 1, 2 < 3 | 4.9** |
| Intell. Words | 4.4 (7.1) | 1.5 (2.7) | 9.5 (9.7) | 10.5** | 1, 2 < 3 | 5.5** |
| NDW | 2.2 (3.3) | 0.8 (1.3) | 1.7 (1.5) | 4.2 | — | 4.1* |
| Total Utterances | 7.3 (3.9) | 7.3 (3.7) | 10.4 (5.5) | 4.3 | — | 2.2 |
| **Non-Word NLS** |  |  |  |  |  |  |
| Non-lexical | 1.8 (1.8) | 2.4 (2.2) | 3.3 (3.8) | 1.8 | — | 1.8 |
| Single-Phoneme | 0.5 (0.4) | 1.1 (1.1) | 0.6 (0.5) | 5.8 | — | 9.5*** |
| Multi-Phoneme | 3.3 (2.5) | 2.8 (2.1) | 4.1 (3.3) | 1.2 | — | 3.4* |

Note. Values are means with standard deviations in parentheses. MLUw = mean length of utterance in words; NDW = number of different words. Kruskal-Wallis tests evaluated group differences; significant tests were followed by FDR-corrected Dunn's post-hoc comparisons. Pairwise labels are 1 = AUT-LI, 2 = DS, and 3 = FXS. Brown-Forsythe tests evaluated variance differences. \* $p < .05$ , \*\* $p < .01$ , \*\*\* $p < .001$ .

Diagnostic groups differed in chronological age, with the AUT-LI group (M = 50.4 months) being significantly older than the DS group (M = 42.1 months) (H = 6.6, p < .05). However, the groups did not differ in mean MSEL non-verbal developmental scores (NVDQ) or various PLS scores (Table 3), suggesting that the differences in chronological age were not accompanied by differences in foundational non-verbal or language ability.

Group differences were observed on NLS measures (Table 4). The FXS group produced significantly higher MLUw, intelligible words, word approximations, and overall intelligibility compared to both the AUT-LI and DS groups. In contrast to the word-based NLS measures, non-word NLS measures showed no significant difference in central tendency.

Brown–Forsythe (F) tests of variance showed significant differences in variance between groups across both PLS measures (AE, GSV, DQ), as well as word (MLUw, word approximations, intelligible words) and non-word (single and multi-phoneme vocalizations) NLS measures.

### Data-Driven Clustering of Expressive Profiles

To explore whether non-word NLS measures provided additional information about the heterogeneity of expressive communication profiles beyond traditional language measures, a two-stage clustering approach consisting of principal component analysis (PCA) followed by partitioning around medoids (PAM) clustering was used. Prior to PCA, candidate NLS measures were screened for redundancy using Spearman correlations (Figure 4A). Several word-based NLS measures were highly correlated, including MLUw, intelligible words, NDW, and percent intelligible speech (r = .90–.99). To avoid entering highly overlapping measures into the PCA, percent intelligible speech was retained because of its clinical interpretability as an index of functional speech clarity, while MLUw, NDW, and intelligible words were excluded from the clustering models. These redundancy reduction rules were applied to both Model A and Model B.

**Figure 4.**
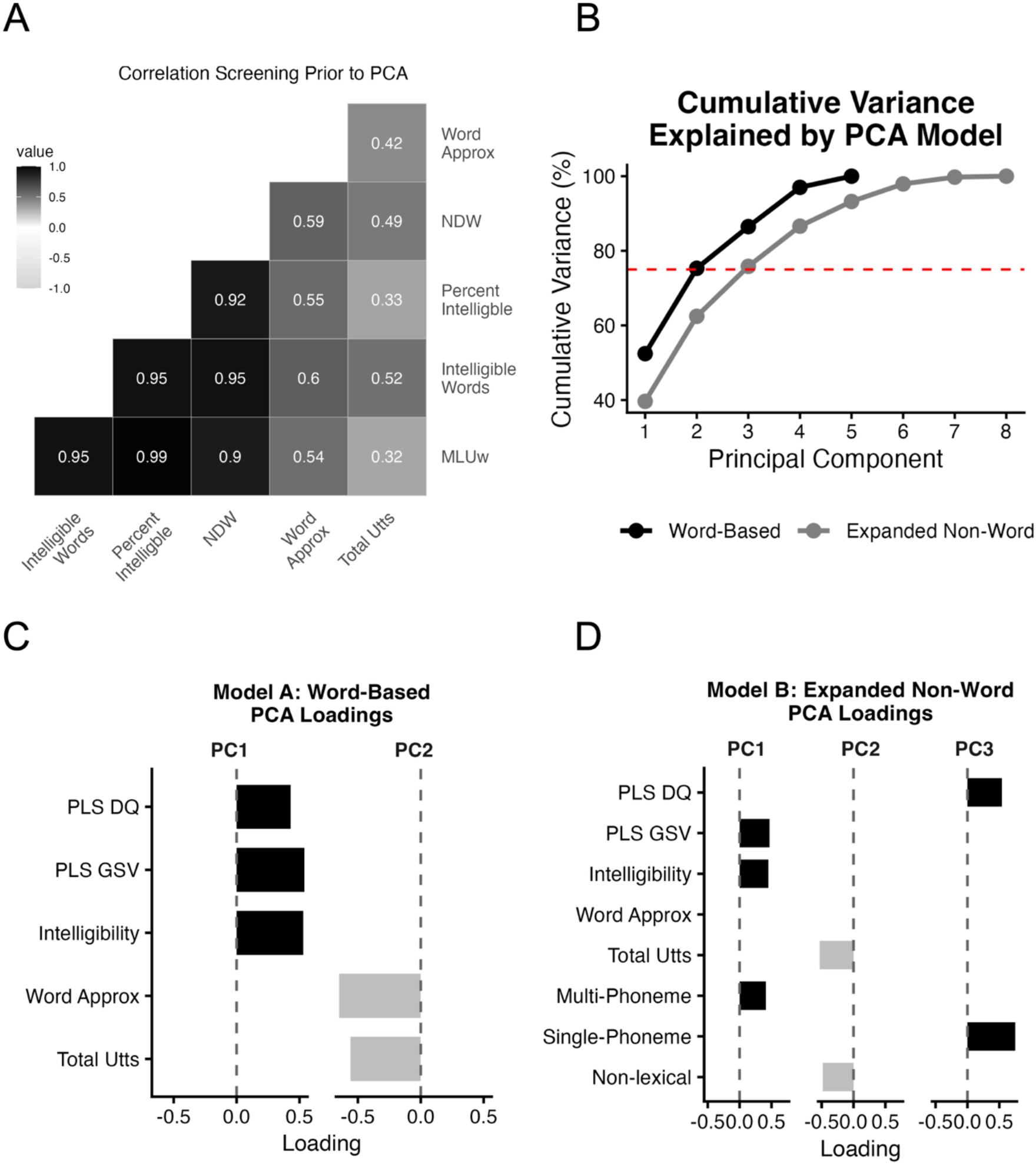
**(A-D**). Principal Component Analysis (PCA) Summary: (A) Spearman correlations used for redundancy screening; color indicates direction and magnitude. (B) cumulative variance explained by each PCA model; dashed line marks the 75% retention threshold. (C-D) PCA loadings ≥ |.40| for retained components in the word-based and expanded nonword models, respectively. PLS = Preschool Language Scales; GSV = growth scale value; DQ = developmental quotient; Tot Utts = total utterances; Word Approx = word approximations.

Using this approach, we compared two exploratory clustering frameworks: Model A serves as a baseline that included standardized language scores (PLS DQ and PLS GSV) and common word-based NLS measures (percent intelligible, total utterances, and word approximations). Model B extended this approach by integrating non-word measures of prelinguistic complexity, stratified into non-lexical sounds, single-phoneme vocalizations, and multi-phoneme vocalizations.

### Principal Component Analysis and Loading Patterns

PCA identified two principal components in Model A and three principal components in Model B, together accounting for approximately 75% of the variance in each model (Figure 4B). Across both models, the first principal component (PC1) showed a consistent pattern of positive loadings for standardized language and intelligibility measures, reflecting a general expressive language dimension characterized by overall expressive output and proficiency (Figure 4C). In Model B, the first principal component included positive loadings for PLS GSV scores, but not PLS DQ, along with intelligibility and multi-phoneme vocalizations, suggesting this component grouped together stronger standardized language performance, greater intelligibility, and more speech-like vocal output (Figure 4D).

The second principal component (PC2) in Model A captures variation in vocal output, with stronger loadings for utterance rate and word approximations. In Model A, the second principal component included strong loadings for total utterances and word approximations, indicating variation in vocal output within the word-based model (Figure 4C). In Model B, individual profiles were further characterized by variation across PC2 and PC3 (Figure 4D). PC2 reflected broader vocal output and non-lexical production, while the third principal component reflected variation related to single-phoneme production and developmental level, as indexed by the PLS developmental quotient. Together, these loading patterns suggest that the inclusion of non-word measures captures additional variation in vocalization profiles that is not represented by word-based measures alone.

### Exploratory Characterization of Communication Profiles

Model A clustering favored a two-cluster solution (silhouette = 0.56, gap statistic = 0.60), which partitioned the sample into a Higher Language, with overall higher performance across word-based metrics, and a Lower Language group with lower performance across the same measures (Figure 5A-B). For Model B, cluster-validity indices showed somewhat stronger support for a two-cluster solution than a three-cluster solution, with average silhouette widths of 0.39 for *k* = 2 and 0.32 for *k* = 3, and gap statistics were 0.42 and 0.41, respectively. However, given the exploratory aim of evaluating whether non-word vocalization measures revealed additional heterogeneity in expressive communication profiles, we examined the three-cluster solution as a descriptive, hypothesis-informed model. This solution suggested three expressive communication profiles: a Higher Language group, characterized by higher standardized language scores and intelligibility; a Low Output group, characterized by reduced vocal production across measures; and a High Output-Low Structure group, characterized by increased vocal production across phonological forms but lower structured language and intelligibility (Figure 5C-D).

**Figure 5.**
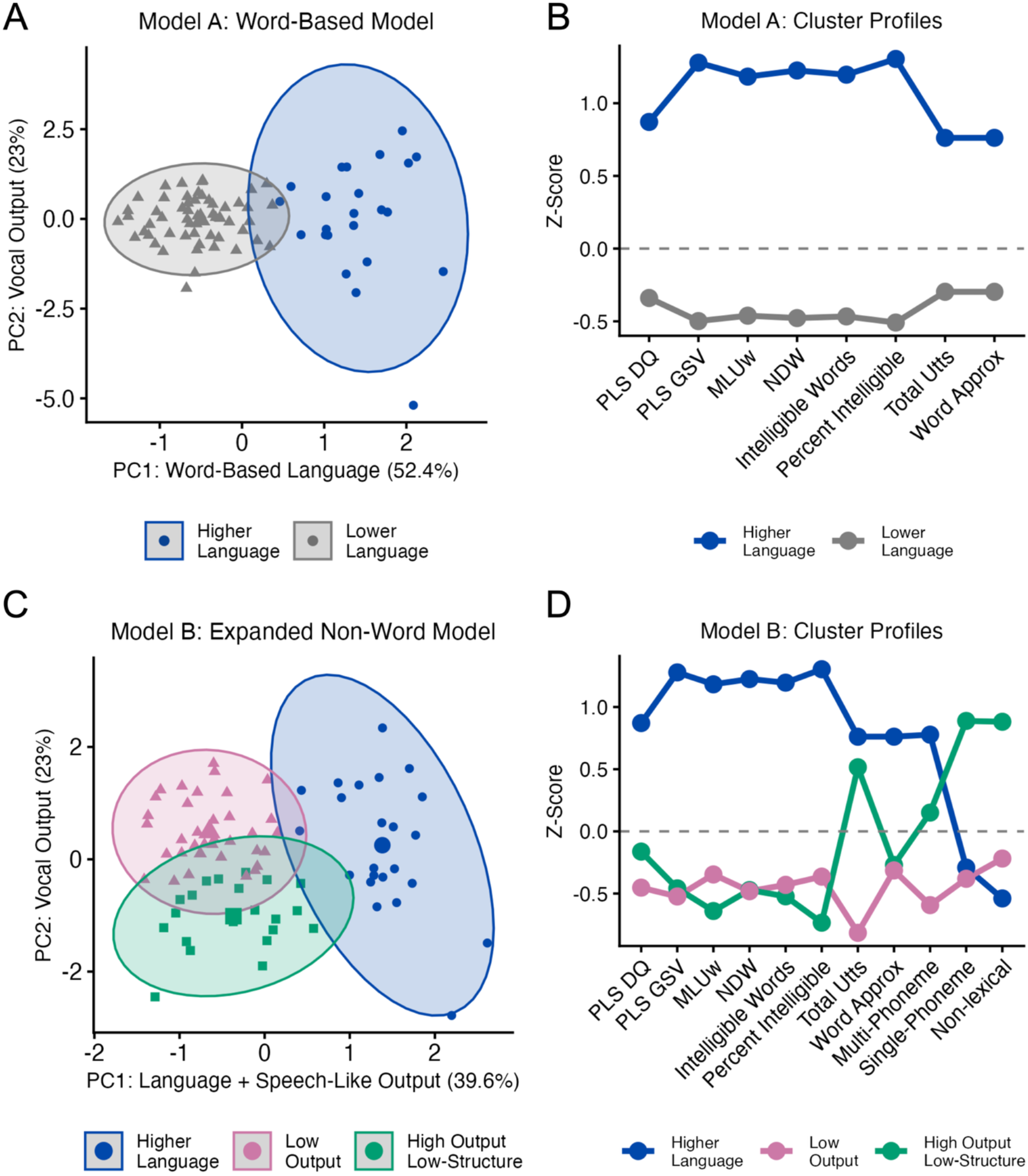
**(A-D).** Cluster solutions and mean communication profiles. PC summaries are based on loadings ≥ |.40|. Model A retained two principal components: PC1 = PLS DQ, PLS GSV, and percent intelligible speech; PC2 = total utterances and word approximations. Model B retained three principal components. Panel 5C cluster solution in the PC1–PC2 space. For Model B, PC1 = PLS GSV, intelligibility, and multi-phoneme vocalizations; PC2 = total utterances and non-lexical vocalizations; PC3 = single-phoneme vocalizations.

### Cluster Characterization and Syndrome-Related Patterns

Finally, we characterized the exploratory three-cluster solution from Model B using external clinical and developmental variables, including chronological age, nonverbal ability, diagnostic group membership, and subsequent monthly change in PLS expressive communication GSV (Figure 6).

**Figure 6.**
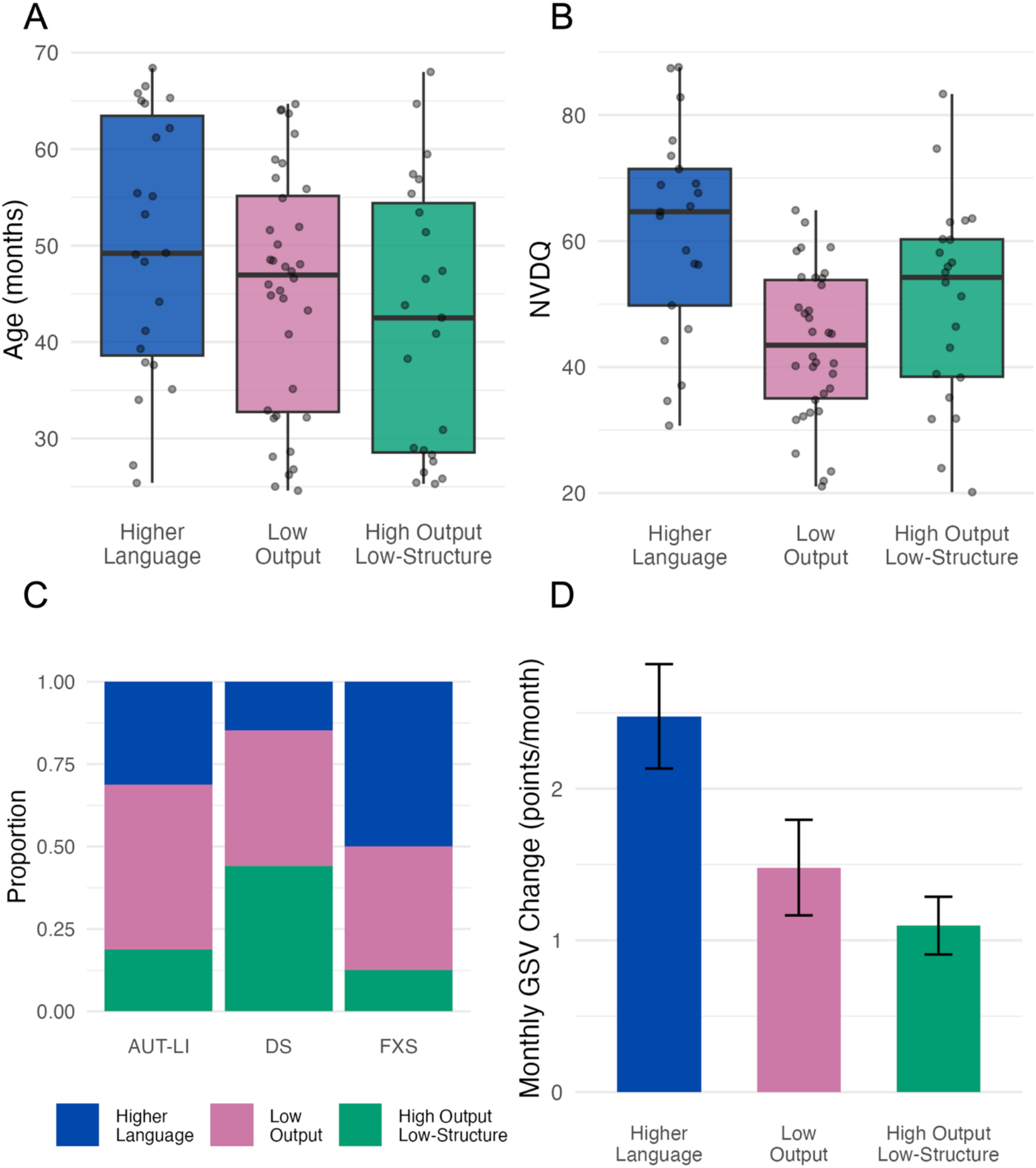
**(A-D).** External characterization of Model B communication profiles: (A) chronological age by profile. (B) nonverbal developmental quotient (NVDQ) by profile. (C) communication profile distribution within diagnostic group. (D) monthly PLS Expressive Communication GSV change by profile among participants with Time Point 2 data (Higher Language: n = 17; Low Output: n = 21; High Output–Low Structure: n = 13). Error bars represent standard errors.

Clusters did not differ significantly in chronological age (F(2, 79) = 2.01, p = .141, but differed in NVDQ, F(2, 74) = 10.14, p < .001. Post-hoc Tukey tests showed that the Higher Language group had higher NVDQ scores than both the Lower Output and High Output-Low Structure groups, which did not differ from one another. Diagnostic group membership was also associated with cluster membership (Fisher’s exact p = .033). Within the AUT-LI group, participants were most commonly classified in the Low Output group (50.0%), whereas DS participants were most commonly classified in the High Output-Low Structure group (44.1%) and FXS participants were most commonly classified in the Higher Language group (50.0%). Standardized residuals indicated that DS was overrepresented in the High Output-Low Structure group and FXS was overrepresented in the Higher Language group (Figure 6C).

Next, we explored whether Time Point 1 communication profiles were associated with subsequent change in PLS expressive communication GSV among participants with Time Point 2 data (n = 51; Higher Language: n = 17; Lower Output: n = 21; High Output-Low Structure: n = 13). This longitudinal subset included children with AUT-LI (n = 18), DS (n = 26), and FXS (n = 7), and analyses were interpreted descriptively because the subset was smaller and diagnostically uneven. Monthly GSV change was calculated as the change in PLS expressive communication GSV divided by months between assessments. Monthly GSV change differed across clusters (χ²(2) = 7.84, p = .020). Pairwise Wilcoxon tests indicated that the Higher Language group showed greater monthly gains than the High Output-Low Structure group after Bonferroni correction (p = .021), whereas the difference between the Higher Language and Lower Output groups was not statistically significant after correction (p = .074). In an adjusted sensitivity analysis with complete covariate data (n = 50), cluster membership remained associated with monthly GSV change after adjusting for baseline PLS GSV, NVDQ, and diagnostic group (F(2, 43) = 3.24, p = .049)).

## Discussion

The current study examined communicative heterogeneity among preschoolers with AUT-LI, FXS, and DS using standardized language measures, word-based NLS measures, and non-word vocalization measures. In the exploratory three-cluster solution in the model that included non-word vocalizations features, children were characterized by three expressive communication profiles: Higher Language, Low Output, and High Output-Low Structure. These profiles reflected differences in standardized language performance, intelligibility, overall vocal output, and vocalization structure, suggesting that non-word vocalization measures may provide descriptive information about expressive communication patterns not captured by diagnostic groups or standardized language scores alone.

### Standardized and natural language measures capture distinct age-related patterns

Expressive language patterns depended on how language ability was measured. Across preschool children with AUT-LI, DS, and FXS, PLS standard scores declined with age, but PLS expressive communication GSVs significantly increased with age. This suggests that while these cohorts show expressive language gains, age-matched expectations are not met. Second, the age-related patterns for both PLS SS and PLS GSV differed across diagnostic groups, suggesting that expressive language development may follow distinct patterns across preschool children with autism, DS, and FXS. Interestingly, many age-related patterns for lexical NLS measures (MLUw, NDW, and intelligibility) increased with age, while the use of non-word single-phoneme vocalizations significantly decreased with age. This may suggest a transition toward more complex multi-phoneme and lexical structures as children age (Marschik et al., 2022). Continuing to explore these cross-group differences may facilitate more sensitive phenotyping and targeted intervention (Hamrick et al., 2025).

Although the clinical groups did not show significant differences on standardized language scores, group differences were observed across both word-based and non-word NLS measures. The children with FXS showed higher expressive productivity across several lexical NLS measures, while non-word vocalization measures revealed differences in variability across clinical groups. These findings suggest that NLS measures capture dimensions of expressive communication that may not be fully reflected in standardized assessment scores alone (Condouris et al., 2003). In particular, combining lexical and non-word NLS measures may provide a more detailed description of functional communication, including differences in vocal output, intelligibility, and emerging speech-like production. These detailed descriptions may be useful for progress monitoring by helping detect not only milestone improvements, but also seemingly small yet meaningful “inch-stone” gains (Hamrick et al., 2025).

### The Value of Non-Word Vocalization Coding

Including non-word vocalization measures expanded the characterization of expressive communication profiles beyond traditional word-based NLS metrics. The primary dimension of overall expressive language ability remained consistent across PCA models; however, the addition of non-lexical, single-phoneme, and multi-phoneme vocalization measures introduced additional dimensions related to broader vocal production and vocalization structure. These added features suggested heterogeneity amongst children with lower language ability, who showed different patterns of vocal output, separating those with generally reduced vocal production from those with higher vocal output but lower structured language and intelligibility. Thus, although exploratory, the expanded Model B feature set suggested that non-word vocalization measures may help characterize clinically meaningful variation among children who show similar profiles on standardized or word-based language measures.

External clinical comparisons provided additional descriptive context for these profiles. The Higher Language group had higher NVDQ scores than the two lower-language groups, whereas the Low Output and High Output-Low Structure groups did not differ significantly from one another in nonverbal developmental quotients. This pattern suggests that differences between the two lower-language profiles may reflect variation in vocal and phonological production rather than broad differences in nonverbal developmental level. However, these exploratory comparisons should be interpreted cautiously given the modest and diagnostically uneven sample.

### Syndrome-Related Patterns in Communication Profiles

Clinical groups were represented across clusters, suggesting that the communication profiles were not simply proxies for diagnostic categories. At the same time, diagnostic group membership was associated with cluster membership (Fisher’s exact *p* = .033), indicating that some profile distributions differed by clinical condition. These findings suggest that expressive language profiles may capture dimensions of heterogeneity that cut across diagnostic labels while still reflecting some diagnosis-associated patterns.

Among children with FXS, the largest proportion of children was classified in the Higher Language group (8/16; 50.0%). This pattern is broadly consistent with prior work showing relative strengths in expressive language and grammatical development in FXS compared with some other neurodevelopmental conditions (Abbeduto et al., 2014; Finestack & Abbeduto, 2010). However, FXS is also associated with substantial within-condition variability in language and cognitive outcomes, which may explain why children with FXS were distributed across multiple profiles (Meng et al., 2021). Co-occurring autism may have further contributed to this heterogeneity. In the current sample, 7 of the 16 children with FXS met criteria for autism and these children were distributed across the Higher Language (3/7), Lower Output (3/7), and High Output-Low Structure (1/7) clusters.

Among children with DS, most were classified in the High Output-Low Structure profile (15/34; 44.1%) or Low Output profile (14/34; 41.2%), with relatively few classified in the Higher Language profile (5/34; 14.7%). This pattern suggests that many children with DS in this sample showed lower structured expressive language, with variability in overall vocal output. This profile may be consistent with prior work documenting persistent challenges in speech intelligibility among children with DS, which may be influenced by anatomical and motor-speech factors including relative marcoglossia, differences in palatal anatomy, and hypotonia affecting the lips, tongue, jaw, and respiratory muscles (Kent & Vorperian, 2013; Hamrick et al., 2025). Although social strengths are often observed in children with DS, co-occurring autism is elevated in this population with an estimated 16-18% of individuals with DS meeting diagnostic criteria (Richards et al., 2015). In the current sample, 1 of 34 children with DS met criteria for autism and was classified in the High Output-Low Structure profile.

Children with ASD-LI were distributed across all three communication profiles, with the largest proportion in the Low Output profile (16/32; 50.0%), followed by the Higher Language profile (10/32; 31.3%), and the High Output-Low Structure profile (6/32; 18.8%). This pattern is consistent with the heterogeneity of expressive language and vocal communication often observed in autistic children with language impairment, including variability in vocal output, intelligibility, and spontaneous communication (DeVeney et al., 2021; Patten et al., 2014). Rather than suggesting a single AUT-LI-specific profile, these findings indicate that NLS-based clustering may help describe expressive communication differences within and across diagnostic categories.

### Limitations and Future Directions

This study contributes to the growing literature on early communication in NDDs by demonstrating the utility of tiered vocalization frameworks applied to natural language sampling. However, several limitations should be noted. First, sample sizes for some diagnostic groups, particularly FXS, were relatively small. This sample size limited our ability to examine communication profiles in children with FXS and DS with co-occurring autism. Future studies with larger and more diverse groups of children will be important in determining whether the communicative patterns identified here remain consistent across broader populations, and whether specific communication profiles emerge in children with genetic disorders and co-occurring autism.

Second, the clustering analyses were exploratory and should be interpreted as a descriptive framework rather than a validated classification system. Although the three-cluster Model B solution was examined to determine whether non-word vocalization measures differentiated children with similar expressive language levels, internal clustering indices also supported a two-cluster solution. The three profiles described here should be interpreted as an exploratory descriptive model. Future studies should examine the stability and reproducibility of these profiles in larger independent samples.

Lastly, age-related findings were based on cross-sectional associations. As a result, these patterns should not be interpreted as individual developmental trajectories. Because the communication profiles were derived from Time Point 1 NLS features, the stability of these vocal profiles over time also cannot yet be determined. Although longitudinal PLS data were available for a subset of participants, repeated NLS data are needed to examine whether profiles such as Low Output or High Output-Low Structure persist across development and whether changes in profile membership relate to later language outcomes, including the likelihood of remaining minimally verbal during the school-age years.

Future studies could also include additional indicators of communication. Behaviors such as gesture use, eye gaze, and caregiver-child interaction patterns may help provide a more complete understanding of how early communication develops and how vocal behaviors interact with other forms of social communication.

Another important direction for future work is improving the feasibility of natural language sampling approaches for use in both research and clinical settings. Prior work suggests that shorter transcription samples may capture vocabulary use reliably in young children. However, more research is needed to determine whether shorter samples can also capture the full range of vocal behaviors in children with neurodevelopmental disorders, particularly for non-word vocalizations. Advances in automated or semi-automated speech processing may also help address this challenge. Continued development of tools that automatically detect and transcribe child vocalizations could reduce the time required for manual transcription while still allowing researchers to capture detailed patterns in early vocal behavior work (McGonigle et al., 2024).

## Conclusions

Together, these findings suggest that combining standardized language measures with word-based and non-word NLS measures can provide a more nuanced characterization of expressive communication in preschoolers with neurodevelopmental disorders. Rather than relying on broader distinctions between “verbal” and “nonverbal,” profile-based characterization may help identify specific barriers to expressive language production. Children with High Output-Low Structure profiles may benefit from interventions that target speech clarity, speech-motor control, and intelligibility. Children with Low Output profiles may benefit from strategies designed to increase overall communication attempts and volubility. Overall, these findings indicate that early communication differences may be better conceptualized as heterogeneous vocal patterns and language profiles rather than a binary distinction between “verbal” and “nonverbal.” Early emerging vocal patterns associated with specific developmental conditions may inform the development of more tailored support strategies.

## Acknowledgments

We thank all the children and families who generously participated in this research. We thank all the research staff and coordinators involved including Meagan Tsou, McKena Geiger, Sophie Hurewitz, Anjali Bose in participant recruitment, data collection, and database administration. This research was supported by the National Institutes of Health (K23DC07983 to CLW, T32DC000038 to AA), the Charles Hood Foundation and the Rosamund Stone Zander Translational Neuroscience Center.

## Data Availability Statement

The datasets generated during and/or analyzed during the current study are available from the corresponding author on reasonable request. The transcription protocol and analysis program are available for download on GitHub: [https://github.com/cwilkinsonlab/nls-transcription/tree/NLS-Transcription-Protocol-and-Pipeline].

## Artificial Intelligence Statement

Artificial Intelligence Statement: ChatGPT (GPT-5.5) was used during manuscript revision to provide language-editing assistance for portions of the manuscript, including assistance with grammar and adherence to word-count limits. Resulting text was revised by the authors prior to inclusion in the manuscript (OpenAI, 2026).

## Author Contributions

Conceptualization: AA, CW

Methodology: AA, CW, KJ, GM, AT

Software: AT, TC

Validation: AA, AT, MN, TC, AM, XH

Formal analysis: AA, GM, AT

Investigation: AA, AM, MN, TC, XH

Writing – original draft: AA, CW

Writing – review & editing: AA, AM, AT, CW, GM, KJ, KP, MN, NB, TC, XH

Visualization: AA, AT

Supervision: CW, KP, NB

Project administration: CW, NB, KP

Funding acquisition: CW, KP, NB

## References

Abbeduto, L., McDuffie, A., & Thurman, A. J. (2014). The fragile X syndrome—Autism comorbidity: What do we really know? Frontiers in Genetics, 5. 10.3389/fgene.2014.00355

Barokova, M., Hassan, S., Lee, C., Xu, M., & Tager-Flusberg, H. (2020). A Comparison of Natural Language Samples Collected From Minimally and Low-Verbal Children and Adolescents With Autism by Parents and Examiners. Journal of Speech, Language, and Hearing Research, 63(12), 4018–4028. 10.1044/2020_JSLHR-20-00343

Barokova, M., La Valle, C., Hassan, S., Lee, C., Xu, M., McKechnie, R., Johnston, E., Krol, M. A., Leano, J., & Tager-Flusberg, H. (2021). Eliciting Language Samples for Analysis (ELSA): A New Protocol for Assessing Expressive Language and Communication in Autism. Autism Research, 14(1), 112–126. 10.1002/aur.2380

Barokova, M., & Tager-Flusberg, H. (2018). Commentary: Measuring Language Change Through Natural Language Samples. Journal of Autism and Developmental Disorders, 50(7), 2287–2306. 10.1007/s10803-018-3628-4

Belardi, K., Watson, L. R., Faldowski, R. A., Hazlett, H., Crais, E., Baranek, G. T., McComish, C., Patten, E., & Oller, D. K. (2017). A Retrospective Video Analysis of Canonical Babbling and Volubility in Infants with Fragile X Syndrome at 9–12 Months of Age. Journal of Autism and Developmental Disorders, 47(4), 1193–1206. 10.1007/s10803-017-3033-4

Brady, N. C., Bruce, S., Goldman, A., Erickson, K., Mineo, B., Ogletree, B. T., Paul, D., Romski, M. A., Sevcik, R., Siegel, E., Schoonover, J., Snell, M., Sylvester, L., & Wilkinson, K. (2016). Communication Services and Supports for Individuals With Severe Disabilities: Guidance for Assessment and Intervention. American Journal on Intellectual and Developmental Disabilities, 121(2), 121–138. 10.1352/1944-7558-121.2.121

Brady, N. C., Kosirog, C., Fleming, K., & Williams, L. (2021). Predicting progress in word learning for children with autism and minimal verbal skills. Journal of Neurodevelopmental Disorders, 13(1), 36. 10.1186/s11689-021-09386-x

CDC. (2025, January 31). Data and Statistics on Fragile X Syndrome. Fragile X Syndrome (FXS). https://www.cdc.gov/fragile-x-syndrome/data/index.html

CDC. (2026, January 30). Developmental Disability Basics. Child Development. https://www.cdc.gov/child-development/about/developmental-disability-basics.html

Chapman, R. (2006). Language learning in Down syndrome: The speech and language profile compared to adolescents with cognitive impairment of unknown origin. Down Syndrome Research and Practice, 10(2), 61–66. 10.3104/reports.306

Condouris, K., Meyer, E., & Tager-Flusberg, H. (2003). The Relationship Between Standardized Measures of Language and Measures of Spontaneous Speech in Children With Autism. American Journal of Speech-Language Pathology, 12(3), 349–358. 10.1044/1058-0360(2003/080)

Costanza-Smith, A. (2010). The Clinical Utility of Language Samples. Perspectives on Language Learning and Education, 17(1), 9–15. 10.1044/lle17.1.9

DeVeney, S. L., Kyvelidou, A., & Mather, P. (2021). A home-based longitudinal study of vocalization behaviors across infants at low and elevated risk of autism. Autism & Developmental Language Impairments, 6, 23969415211057658. 10.1177/23969415211057658

Eigsti, I.-M., De Marchena, A. B., Schuh, J. M., & Kelley, E. (2011). Language acquisition in autism spectrum disorders: A developmental review. Research in Autism Spectrum Disorders, 5(2), 681–691. 10.1016/j.rasd.2010.09.001

Fidler, D. J. (2005). The Emerging Down Syndrome Behavioral Phenotype in Early Childhood: Implications for Practice. Infants & Young Children, 18(2), 86–103. 10.1097/00001163-200504000-00003

Finestack, L. H., & Abbeduto, L. (2010). Expressive Language Profiles of Verbally Expressive Adolescents and Young Adults With Down Syndrome or Fragile X Syndrome. Journal of Speech, Language, and Hearing Research, 53(5), 1334–1348. 10.1044/1092-4388(2010/09-0125)

Finestack, L. H., Richmond, E. K., & Abbeduto, L. (2009). Language Development in Individuals With Fragile X Syndrome. Topics in Language Disorders, 29(2), 133–148. 10.1097/TLD.0b013e3181a72016

Gary, T., & Wallace, S. (2020). From the Editors: Language Sample Analysis: New and Neglected Clinical Applications. Topics in Language Disorders, 40(2), 131–131. 10.1097/TLD.0000000000000216

Hamrick, L. R., Boorom, O., Estrada, K., Brady, N., & Kelleher, B. (2025). Prelinguistic Communication Complexity of Children With Neurogenetic Syndromes. Journal of Speech, Language, and Hearing Research, 68(8), 3938–3955. 10.1044/2025_JSLHR-24-00477

Heilmann, J., & Miller, J. F. (2023). Systematic Analysis of Language Transcripts Solutions: A Tutorial. Perspectives of the ASHA Special Interest Groups, 8(1), 1–18. 10.1044/2022_PERSP-22-00148

Hessl, D., Nguyen, D. V., Green, C., Chavez, A., Tassone, F., Hagerman, R. J., Senturk, D., Schneider, A., Lightbody, A., Reiss, A. L., & Hall, S. (2009). A solution to limitations of cognitive testing in children with intellectual disabilities: The case of fragile X syndrome. Journal of Neurodevelopmental Disorders, 1(1), 33–45. 10.1007/s11689-008-9001-8

Iverson, J. M., & Wozniak, R. H. (2007). Variation in Vocal-Motor Development in Infant Siblings of Children with Autism. Journal of Autism and Developmental Disorders, 37(1), 158–170. 10.1007/s10803-006-0339-z

Jhang, Y., & Oller, D. K. (2017). Emergence of Functional Flexibility in Infant Vocalizations of the First 3 Months. Frontiers in Psychology, 8. 10.3389/fpsyg.2017.00300

Johnson, K. T., Narain, J., Quatieri, T., Maes, P., & Picard, R. W. (2023). ReCaNVo: A database of real-world communicative and affective nonverbal vocalizations. Scientific Data, 10(1), 1–13. https://www.nature.com/articles/s41597-023-02405-7

Kasari, C., Brady, N., Lord, C., & Tager-Flusberg, H. (2013). Assessing the Minimally Verbal School-Aged Child With Autism Spectrum Disorder. Autism Research, 6(6), 479–493. 10.1002/aur.1334

Kaufmann, W. E., Kidd, S. A., Andrews, H. F., Budimirovic, D. B., Esler, A., Haas-Givler, B., Stackhouse, T., Riley, C., Peacock, G., Sherman, S. L., Brown, W. T., & Berry-Kravis, E. (2017). Autism Spectrum Disorder in Fragile X Syndrome: Cooccurring Conditions and Current Treatment. Pediatrics, 139(Supplement_3), S194–S206. 10.1542/peds.2016-1159F

Kent, R. D., & Vorperian, H. K. (2013). Speech impairment in Down syndrome: A review. Journal of Speech, Language, and Hearing Research, 56(1), 178–210. 10.1044/1092-4388(2012/12-0148)

La Valle, C., Shen, L., Shih, W., Kasari, C., Shire, S., Lord, C., & Tager-Flusberg, H. (2024). Does Gestural Communication Influence Later Spoken Language Ability in Minimally Verbal Autistic Children? Journal of Speech, Language, and Hearing Research, 67(7), 2283–2296. 10.1044/2024_JSLHR-23-00433

La Valle, C., Plesa-Skwerer, D., & Tager-Flusberg, H. (2020). Comparing the pragmatic speech profiles of minimally verbal and verbally fluent individuals with autism spectrum disorder. Journal of autism and developmental disorders, 50(10), 3699–3713.

Long, H. L., & Hustad, K. C. (2023). Marginal and Canonical Babbling in 10 Infants at Risk for Cerebral Palsy. American journal of speech-language pathology, 32(4S), 1835–1849. 10.1044/2022_AJSLP-22-00165

Lord, C., Brugha, T. S., Charman, T., Cusack, J., Dumas, G., Frazier, T., Jones, E. J. H., Jones, R. M., Pickles, A., State, M. W., Taylor, J. L., & Veenstra-VanderWeele, J. (2020). Autism spectrum disorder. Nature Reviews Disease Primers, 6(1), 5. 10.1038/s41572-019-0138-4

Luyster, R. J., Kadlec, M. B., Carter, A., & Tager-Flusberg, H. (2008). Language Assessment and Development in Toddlers with Autism Spectrum Disorders. Journal of Autism and Developmental Disorders, 38(8), 1426–1438. 10.1007/s10803-007-0510-1

Lynch, M. P., Oller, D. K., Steffens, M. L., & Buder, E. H. (1995). Phrasing in prelinguistic vocalizations. Developmental Psychobiology, 28(1), 3–25. 10.1002/dev.420280103

Maes, P., Weyland, M., & Kissine, M. (2022). Describing (pre)linguistic oral productions in 3- to 5-year-old autistic children: A cluster analysis. Autism, 27(4), 967–982. 10.1177/13623613221122663

Marschik, P. B., Widmann, C. A. A., Lang, S., Kulvicius, T., Boterberg, S., Nielsen-Saines, K., Bölte, S., Esposito, G., Nordahl-Hansen, A., Roeyers, H., Wörgötter, F., Einspieler, C., Poustka, L., & Zhang, D. (2022). Emerging Verbal Functions in Early Infancy: Lessons from Observational and Computational Approaches on Typical Development and Neurodevelopmental Disorders. Advances in Neurodevelopmental Disorders, 6(4), 369–388. 10.1007/s41252-022-00300-7

Narain, J., Johnson, K. T., O’Brien, A., Wofford, P., Maes, P., & Picard, R. (2020a). Nonverbal vocalizations as speech: Characterizing natural-environment audio from nonverbal individuals with autism. Proceedings of Laughter and Other Non-Verbal Vocalisations Workshop. https://biecoll.ub.uni-bielefeld.de/index.php/lw2020/article/download/923/947

Narain, J., Johnson, K. T., Ferguson, C., O’Brien, A., Talkar, T., Weninger, Y. Z., Wofford, P., Quatieri, T., Picard, R., & Maes, P. (2020b). Personalized modeling of real-world vocalizations from nonverbal individuals. Proceedings of the 2020 International Conference on Multimodal Interaction, 665–669. 10.1145/3382507.3418854

OpenAI. (2026). ChatGPT (GPT-5.5) [Large language model]. https://chatgpt.com/

McGonigle, E., VanDam, M., Wilkinson, C., & Johnson, K. T. (2024). Benchmarking Automatic Speech Recognition Technology for Natural Language Samples of Children With and Without Developmental Delays. 2024 46th Annual International Conference of the IEEE Engineering in Medicine and Biology Society (EMBC), 1–5. 10.1109/EMBC53108.2024.10782773

Meng, L., Kaufmann, W. E., Frye, R. E., Ong, K., Kaminski, J. W., Velinov, M., & Berry-Kravis, E. (2021). The association between mosaicism type and cognitive and behavioral functioning among males with fragile X syndrome. American Journal of Medical Genetics Part A. 10.1002/ajmg.a.62594

Miller, J., & Iglesias, A. (2012). Systematic Analysis of Language Transcripts (SALT), Research Version [Computer software]. SALT Software, LLC.

Mullen, E. M. (1995). Mullen Scales of Early Learning. Pearson.

Nathani, S., Ertmer, D. J., & Stark, R. E. (2006). Assessing vocal development in infants and toddlers. Clinical Linguistics & Phonetics, 20(5), 351–369. 10.1080/02699200500211451

Oller, D. K., Buder, E. H., Ramsdell, H. L., Warlaumont, A. S., Chorna, L., & Bakeman, R. (2013). Functional flexibility of infant vocalization and the emergence of language. Proceedings of the National Academy of Sciences, 110(16), 6318–6323. 10.1073/pnas.1300337110

Oller, D. K., & Kent, R. D. (2001). The Emergence of the Speech Capacity. The Journal of the Acoustical Society of America, 110(3), 1237–1238. 10.1121/1.1388001

Patten, E., Belardi, K., Baranek, G. T., Watson, L. R., Labban, J. D., & Oller, D. K. (2014). Vocal Patterns in Infants with Autism Spectrum Disorder: Canonical Babbling Status and Vocalization Frequency. Journal of Autism and Developmental Disorders, 44(10), 2413–2428. 10.1007/s10803-014-2047-4

Pavelko, S. L., Owens, R. E., Ireland, M., & Hahs-Vaughn, D. L. (2016). Use of Language Sample Analysis by School-Based SLPs: Results of a Nationwide Survey. Language, Speech, and Hearing Services in Schools, 47(3), 246–258. 10.1044/2016_LSHSS-15-0044

Peña, E. D., Gillam, R. B., & Bedore, L. M. (2014). Dynamic Assessment of Narrative Ability in English Accurately Identifies Language Impairment in English Language Learners. Journal of Speech, Language, and Hearing Research, 57(6), 2208–2220. 10.1044/2014_JSLHR-L-13-0151

Plate, S. N. (2025). The state of natural language sampling in autism research: A scoping review. Autism & Developmental Language Impairments, 10, 1–22.

Richards, C., Jones, C., Groves, L., Moss, J., & Oliver, C. (2015). Prevalence of autism spectrum disorder phenomenology in genetic disorders: A systematic review and meta-analysis. The Lancet Psychiatry, 2(10), 909–916. 10.1016/S2215-0366(15)00376-4

Roberts, J. E., Bradshaw, J., Will, E., Hogan, A. L., McQuillin, S., & Hills, K. (2020). Emergence and rate of autism in fragile X syndrome across the first years of life. Development and Psychopathology, 32(4), 1335–1352. 10.1017/S0954579420000942

Roche, L., Zhang, D., Bartl-Pokorny, K. D., Pokorny, F. B., Schuller, B. W., Esposito, G., Bölte, S., Roeyers, H., Poustka, L., Gugatschka, M., Waddington, H., Vollmann, R., Einspieler, C., & Marschik, P. B. (2018). Early Vocal Development in Autism Spectrum Disorder, Rett Syndrome, and Fragile X Syndrome: Insights from Studies Using Retrospective Video Analysis. Advances in Neurodevelopmental Disorders, 2(1), 49–61. 10.1007/s41252-017-0051-3

Spaulding, T. J., Plante, E., & Farinella, K. A. (2006). Eligibility Criteria for Language Impairment: Is the Low End of Normal Always Appropriate? Language, Speech, and Hearing Services in Schools, 37(1), 61–72. 10.1044/0161-1461(2006/007)

Tager-Flusberg, H., & Kasari, C. (2013). Minimally Verbal School-Aged Children with Autism Spectrum Disorder: The Neglected End of the Spectrum. Autism Research, 6(6), 468–478. 10.1002/aur.1329

Tager-Flusberg, H., Paul, R., & Lord, C. (2005). Language and Communication in Autism. In Handbook of Autism and Pervasive Developmental Disorders (Vol. 1).

Tager-Flusberg, H., Rogers, S., Cooper, J., Landa, R., Lord, C., Paul, R., Rice, M., Stoel-Gammon, C., Wetherby, A., & Yoder, P. (2009). Defining Spoken Language Benchmarks and Selecting Measures of Expressive Language Development for Young Children With Autism Spectrum Disorders. *Journal of Speech*, Language, and Hearing Research, 52(3), 643–652. 10.1044/1092-4388(2009/08-0136)

Thurm, A., Manwaring, S. S., Swineford, L., & Farmer, C. (2015). Longitudinal study of symptom severity and language in minimally verbal children with autism. Journal of Child Psychology and Psychiatry, 56(1), 97–104. 10.1111/jcpp.12285

Thurman, A. J., Bullard, L., Kelly, L., Wong, C., Nguyen, V., Esbensen, A. J., Bekins, J., Schworer, E. K., Fidler, D. J., Daunhauer, L. A., Mervis, C. B., Pitts, C. H., Becerra, A. M., & Abbeduto, L. (2022). Defining Expressive Language Benchmarks for Children with Down Syndrome. Brain sciences, 12(6), 743. 10.3390/brainsci12060743

Warlaumont, A. S., Richards, J. A., Gilkerson, J., & Oller, D. K. (2014). A Social Feedback Loop for Speech Development and Its Reduction in Autism. Psychological Science, 25(7), 1314–1324. 10.1177/0956797614531023

Woynaroski, T., Watson, L., Gardner, E., Newsom, C. R., Keceli-Kaysili, B., & Yoder, P. J. (2016). Early predictors of growth in diversity of key consonants used in communication in initially preverbal children with autism spectrum disorder. Journal of Autism and Developmental Disorders, 46(3), 1013–1024. 10.1007/s10803-015-2647-7

Zheng, S., Hume, K. A., Able, H., Bishop, S. L., & Boyd, B. A. (2020). Exploring Developmental and Behavioral Heterogeneity among Preschoolers with ASD: A Cluster Analysis on Principal Components. Autism Research, 13(5), 796–809. 10.1002/aur.2263

Zimmerman, Irla Lee, Steiner, Violette G., & Pond, Roberta E. (2011). Preschool Language Scales, Fifth Edition (PLS-5) (5th ed.). Pearson.

